# Virological assessment of hospitalized cases of coronavirus disease 2019

**DOI:** 10.1101/2020.03.05.20030502

**Authors:** Roman Wölfel, Victor M. Corman, Wolfgang Guggemos, Michael Seilmaier, Sabine Zange, Marcel A. Müller, Daniela Niemeyer, Terence C. Jones Kelly, Patrick Vollmar, Camilla Rothe, Michael Hoelscher, Tobias Bleicker, Sebastian Brünink, Julia Schneider, Rosina Ehmann, Katrin Zwirglmaier, Christian Drosten, Clemens Wendtner

## Abstract

Coronavirus disease 2019 (COVID-19) is an acute respiratory tract infection that emerged in late 2019^1,2^. Initial outbreaks in China involved 13.8% cases with severe-, and 6.1% with critical courses^3^. This severe presentation corresponds to the usage of a virus receptor that is expressed predominantly in the lung^2,4^. By causing an early onset of severe symptoms, this same receptor tropism is thought to have determined pathogenicity but also aided the control of severe acute respiratory syndrome (SARS) in 2003^5^. However, there are reports of COVID-19 cases with mild upper respiratory tract symptoms, suggesting a potential for pre- or oligosymptomatic transmission^6-8^. There is an urgent need for information on body site - specific virus replication, immunity, and infectivity. Here we provide a detailed virological analysis of nine cases, providing proof of active virus replication in upper respiratory tract tissues. Pharyngeal virus shedding was very high during the first week of symptoms (peak at 7.11 × 10^8^ RNA copies per throat swab, day 4). Infectious virus was readily isolated from throat- and lung-derived samples, but not from stool samples in spite of high virus RNA concentration. Blood and urine never yielded virus. Active replication in the throat was confirmed by viral replicative RNA intermediates in throat samples. Sequence-distinct virus populations were consistently detected in throat- and lung samples of one same patient. Shedding of viral RNA from sputum outlasted the end of symptoms. Seroconversion occurred after 6-12 days, but was not followed by a rapid decline of viral loads. COVID-19 can present as a mild upper respiratory tract illness. Active virus replication in the upper respiratory tract puts prospects of COVID-19 containment in perspective.

---

There is a close genetic relatedness between severe acute respiratory syndrome coronavirus (SARS-CoV) and the causative agent of COVID-19, SARS-CoV-2. The predominant expression of ACE2 in the lower respiratory tract is believed to have determined the natural history of SARS as a lower respiratory tract infection. Whereas positive SARS-CoV-2 detection in clinical specimens from the upper respiratory tract has been described^**9**^, these observations do not address principal differences between SARS and COVID-19 in terms of clinical pathology. The here-studied patients were enrolled because they acquired their infections upon known close contact to an index case, thereby avoiding representational biases due to symptom-based case definitions. All patients were treated in a single hospital in Munich, Germany. Virological testing was done by two closely-collaborating laboratories using the same standards of technology for RT-PCR and virus isolation, confirming each other’s results based on almost all individual samples. Due to extremely high congruence of results, all data are presented together. Only the serological data are based on results from one laboratory. The patients are part of a larger cluster of epidemiologically-linked cases that occurred after January 23^rd^, 2020 in Munich, Germany, as discovered on January 27^th^ (Böhmer et al., accompanying manuscript). The present study uses samples taken during the clinical course in the hospital, as well as from initial diagnostic testing before admission. In cases when this initial diagnostic testing was done by other laboratories, the original samples were retrieved and re-tested under the rigorous quality standards of the present study.

## RT-PCR sensitivity, sites of replication, and correlates of infectivity based on aggregated data

To first understand whether the described clinical presentations are solely caused by SARS-CoV-2 infection, samples from all patients were tested against a panel of typical agents of respiratory viral infection, including HCoV-HKU1, -OC43, -NL63, -229E; Influenza virus A and B, Rhinovirus, Enterovirus, Respiratory syncytial virus, Human Parainfluenza virus 1-4, Human metapneumovirus, Adenovirus, and Human bocavirus. Interestingly, no co-infection was detected in any patient.

All patients were initially diagnosed by RT-PCR^10^ from oro- or nasopharyngeal swab specimens. Both specimen types were collected over the whole clinical course in all patients. There were no discernible differences in viral loads or detection rates when comparing naso-vs. oropharyngeal swabs (**Figure 1B**). The earliest swabs were taken on day 1 of symptoms, with symptoms often being very mild or prodromal. All swabs from all patients taken between days 1 and 5 tested positive. The average virus RNA load was 6.76×10^5^ copies per whole swab until day 5 (maximum, 7.11×10^8^ copies/swab). Swab samples taken after day 5 had an average viral load of 5.13×10^3^ copies per swab and a detection rate of 45.95%. The last swab sample was taken on day 22 post-onset. Average viral load in sputum was 1.18 × 10^6^ copies per mL (maximum, 6.65×10^8^ copies per mL).

**Figure 1.**
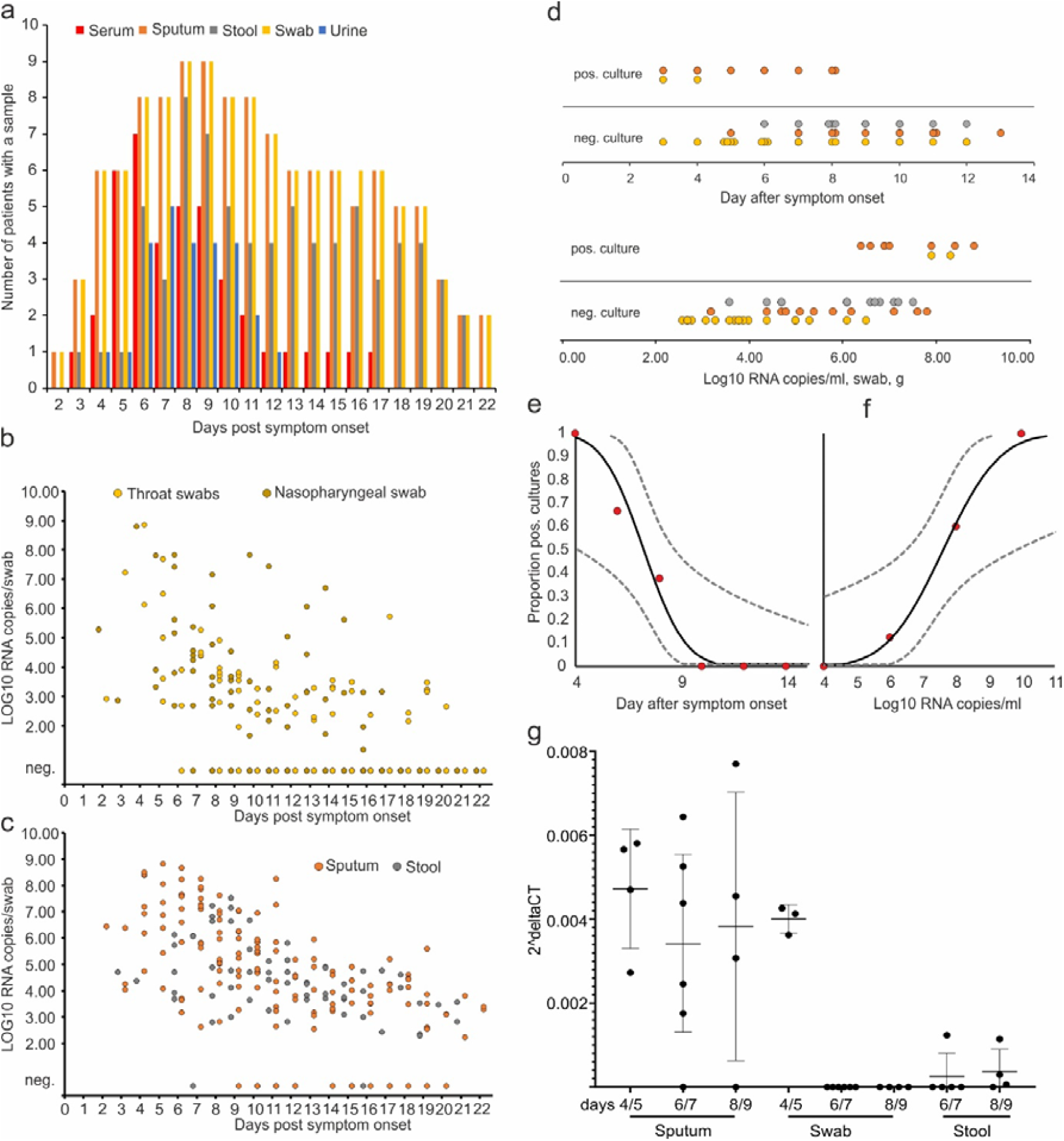
Hallmarks of viral shedding in aggregated samples. A, samples and sample types per day. B, viral RNA concentrations in upper respiratory tract samples. C, viral RNA concentrations in sputum and stool samples. D, virus isolation success dependent on day post-onset. E, virus isolation success dependent on viral load. F and G, projected virus isolation success based on probit distributions. The inner lines are probit curves (dose-response rule). The outer dotted lines are 95% CI. For less than 5% isolation success, the estimated day was 9.78 (95% CI: 8.45-21.78) days post-onset and the estimated RNA concentration for less than 5% isolation success was estimate to be 6.51 Log10 RNA/ml (95% CI:-4,11-5.40). H, Subgenomic viral RNA transcripts in relation to viral genomic RNA.

Because swab samples had limited sensitivity for initial diagnosis of cases of SARS^13,14^, we analyzed the first paired swab and sputum samples taken on the same occasion from seven patients (**Table 1**). All samples were taken between 2 and 4 days post-onset. In two cases, swab samples had clearly higher virus concentrations than sputum samples, as indicated by a difference greater than 3 in threshold cycle (Ct) value. The opposite was true in two others cases, while the other 5 cases had similar concentrations in both sample types. None of 27 urine samples and none of 31 serum samples were tested positive for SARS-CoV2 RNA.

**Table 1.** Single nucleotide polymorphism in clinical samples from case #4

| Day p.o. | 5 | 6 | 7 | 8 | 9 | 10 | 11 |
| --- | --- | --- | --- | --- | --- | --- | --- |
| Swab | A |  | A |  |  |  |  |
| Sputum |  | G | G | G | G>A |  |  |
| Stool |  |  | G>A | A=G | A=G | G>A | A |

To understand infectivity, live virus isolation was attempted on multiple occasions from clinical samples (**Figure 1 D**). Whereas virus was readily isolated during the first week of symptoms from a significant fraction of samples (16.66% in swabs, 83.33% in sputum samples), no isolates were obtained from samples taken after day 8 in spite of ongoing high viral loads.

Virus isolation from stool samples was never successful, irrespective of viral RNA concentration, based on a total of 13 samples taken between days six to twelve from four patients. Virus isolation success also depended on viral load: samples containing <10^6^ copies/mL (or copies per sample) never yielded an isolate. For swab and sputum, interpolation based on a probit model was done to obtain laboratory-based infectivity criteria for discharge of patients (**Figures 1 E, F**).

High viral loads and successful isolation from early throat swabs suggested potential virus replication in upper respiratory tract tissues. To obtain proof of active virus replication in absence of histopathology, we conducted RT-PCR tests to identify viral subgenomic messenger RNAs (sgRNA) directly in clinical samples. Viral sgRNA is only transcribed in infected cells and is not packaged into virions, therefore indicating the presence of actively-infected cells in samples. Viral sgRNA was compared against viral genomic RNA in the same sample. In sputum samples taken on days 4/5, 6/7, and 8/9, a time in which active replication in sputum was obvious in all patients as per longitudinal viral load courses (see below), mean normalized sgRNA per genome ratios were ∼0.4% (**Figure 1G**). Swabs taken up to day 5 were in the same range, while no sgRNA was detectable in swabs thereafter. Together, these data indicate active replication of SARS-CoV-2 in the throat during the first 5 days after symptoms onset. No, or only minimal, indication of replication in stool was obtained by the same method (**Figure 1G**).

During our study we sequenced full virus genomes from all patients. A G6446A exchange was first detected in one patient and later transmitted to other patients in the cluster (Böhmer, accompanying manuscript). In the first patient, this mutation was found in a throat swab while a sputum sample from the same day still showed the original allele, 6446G. The SNP was analyzed by RT-PCR and Sanger sequencing in all sequential samples available from that patient (**Table 1**). The presence of separate genotypes in throat swabs and sputum strongly supported our suspicion of independent virus replication in the throat, rather than passive shedding there from the lung.

## Virus shedding, antibody response, and clinical correlation in individual courses

Daily measurements of viral load in sputum, pharyngeal swabs, and stool are summarized in **Figure 2**. In general, viral RNA concentrations were very high in initial samples. In all patients except one, throat swab RNA concentrations seemed to be already on the decline at the time of first presentation. Sputum RNA concentrations declined more slowly, with a peak during the first week of symptoms in three of eight patients. Stool RNA concentrations were also high. Courses of viral RNA concentration in stool seemed to reflect courses in sputum in many cases (e.g., **Figure 2 A, B, C**). In only one case, independent replication in the intestinal tract seemed obvious from the course of stool RNA excretion (**Figure 2 D**). Whereas symptoms mostly waned until the end of the first week (**Table 2**), viral RNA remained detectable in throat swabs well into the second week. Stool and sputum samples remained RNA-positive over even longer periods, in spite of full resolution of symptoms.

**Table 2.** Clinical characteristics of all patients

| PATIENT ID | COMORBIDITY | INITIAL SYMPTOMS | ANC/μL | ALC/μL | CRP (MG/L) | LDH (U/L) |
| --- | --- | --- | --- | --- | --- | --- |
| #1 | hypothyroidism | cough, fever, diarrhea | 4870 | 1900 | 46 | 197 |
| #2 | none | sinusitis, cephalgia, cough, | 3040 | 1200 | 4.9 | 182 |
| #3 | COPD | arthralgia, sinusitis, cough, | 5040 | 2600 | 1.3 | 191 |
| #4 | none | otitis, rhinitis, | 2420 | 2220 | 5.9 | 149 |
| #7 | hyper-cholesterinemia | rhinitis, cough, | 4690 | 900 | 4.9 | 209 |
| #8 | none | sinusitis, cough | 2500 | 1600 | 1.7 | 203 |
| #10 | none | sinusitis, cough, | 2350 | 700 | 7.8 | 220 |
| #14 | none | fever, cough, diarrhea | 5040 | 1500 | 9.8 | 220 |
| #16 | none | none | 4620 | 900 | 0.5 | 201 |

Abbreviations: ANC = absolute neutrophil count, ALC = absolute lymphocyte count, CRP = C-reactive protein, LDH = lactate dehydrogenase, M = male, F = female

**Figure 2.**
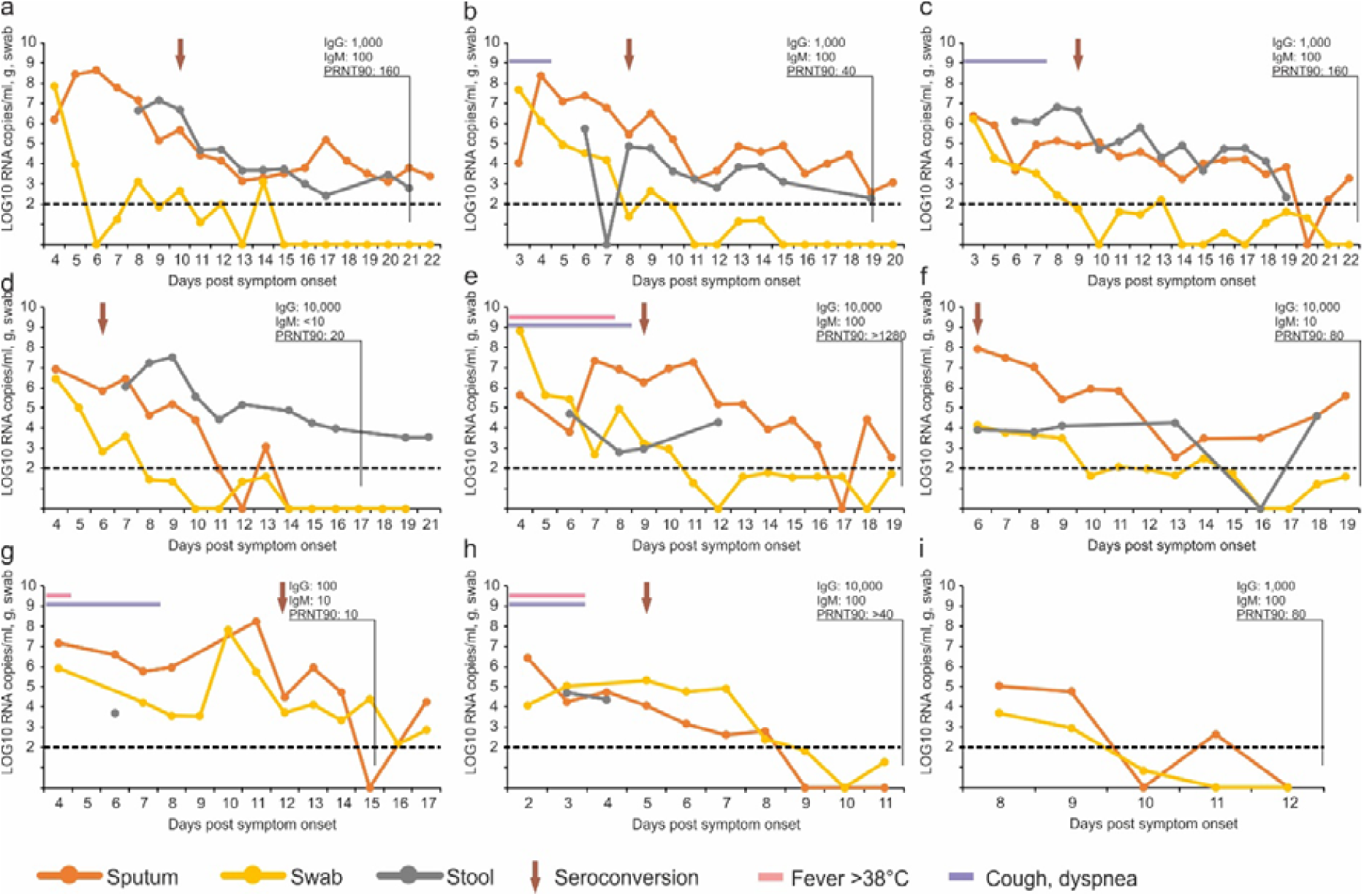
Viral load kinetics, seroconversion and clinical observations in individual cases. Panels A to I correspond to cases #1, #2, #3, #4, #7, #8, #10, #14, and #16 in Böhmer et al.(accompanying manuscript). Dotted lines, limit of quantification

All cases had comparatively mild courses (**Table 2**). The two patients who showed some signs of pneumonia were the only cases where sputum viral loads showed a late and high peak around day 10/11, whereas sputum viral loads were on the decline by this time in all other patients (**Figure 2 F,G**).

Seroconversion was detected by IgG and IgM immunofluorescence using cells expressing the spike protein of SARS-CoV-2 and a virus neutralization assay using SARS-CoV-2 (**Table 3**). In early sera, taken between day 3 and 6, none of the patients showed detectable antibody. The patients monitored long enough to yield a serum sample after two weeks all showed neutralizing antibodies, the titer levels of which did not suggest any correlation with clinical courses. Of note, case #4, with the lowest virus neutralization titer at end of week 2, seemed to shed virus from stool over prolonged time (**Figure 2 D**). Results on differential recombinant immunofluorescence assay indicated no significant rise in titer against the four endemic human Coronaviruses (**Table S1**).

**Table 3.**
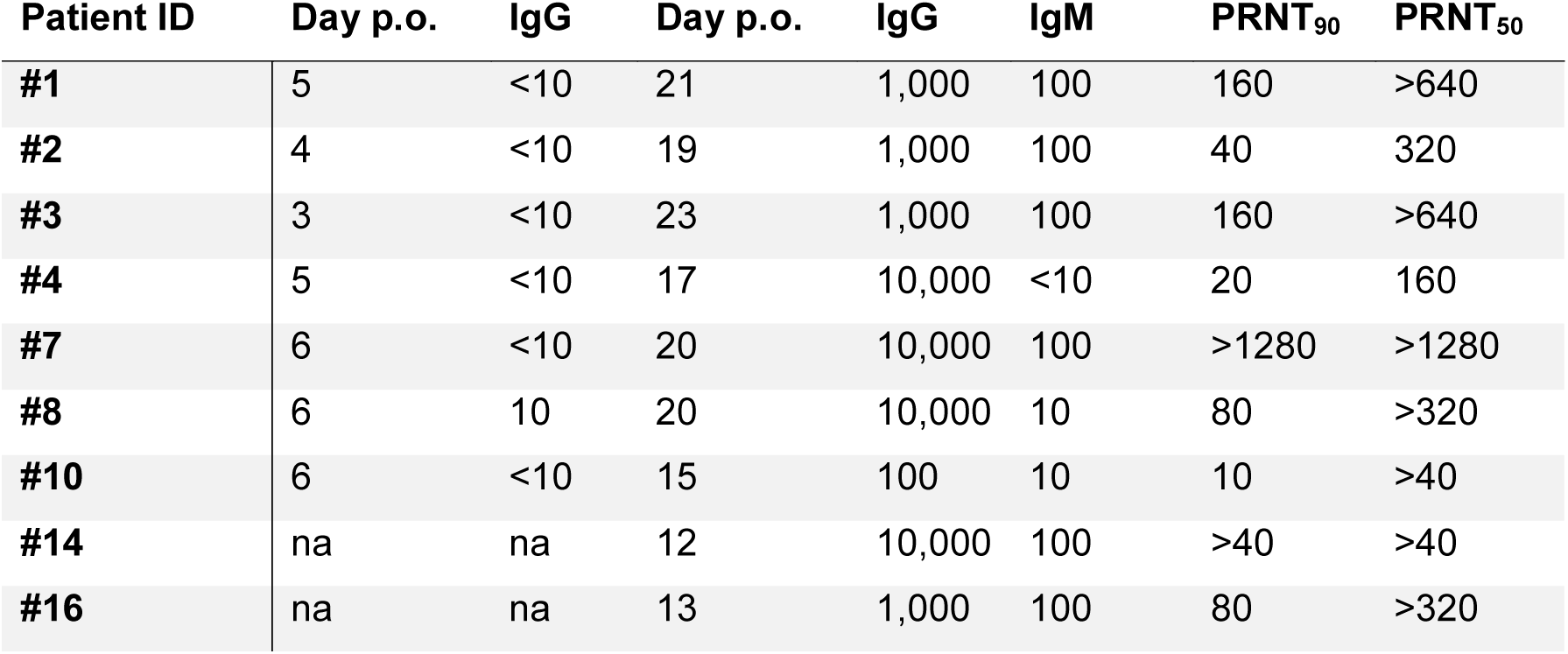
IgG and IgM immunofluorescence titers against SARS-CoV-2 of all patients

| Patient ID | Day p.o. | IgG | Day p.o. | IgG | IgM | PRNT<sub>90</sub> | PRNT<sub>50</sub> |
| --- | --- | --- | --- | --- | --- | --- | --- |
| #1 | 5 | <10 | 21 | 1,000 | 100 | 160 | >640 |
| #2 | 4 | <10 | 19 | 1,000 | 100 | 40 | 320 |
| #3 | 3 | <10 | 23 | 1,000 | 100 | 160 | >640 |
| #4 | 5 | <10 | 17 | 10,000 | <10 | 20 | 160 |
| #7 | 6 | <10 | 20 | 10,000 | 100 | >1280 | >1280 |
| #8 | 6 | 10 | 20 | 10,000 | 10 | 80 | >320 |
| #10 | 6 | <10 | 15 | 100 | 10 | 10 | >40 |
| #14 | na | na | 12 | 10,000 | 100 | >40 | >40 |
| #16 | na | na | 13 | 1,000 | 100 | 80 | >320 |

## Conclusions

The clinical courses in subjects under study were mild, all being young- to middle-aged professionals without significant underlying disease. Apart from one patient, all cases were first tested when symptoms were still mild or in the prodromal stage, a period in which most patients would present once there is general awareness of a circulating pandemic disease^5^. Diagnostic testing suggests that simple throat swabs will provide sufficient sensitivity at this stage of infection. This is in stark contrast to SARS. For instance, only 38 of 98 nasal or nasopharyngeal swab samples tested positive by RT-PCR in SARS patients in Hong Kong^15^. Also, viral load differed considerably. In SARS, it took 7 to 10 days after onset until peak RNA concentrations (of up to 5×10^5^ copies per swab) were reached^13,14^. In the present study, peak concentrations were reached before day 5, and were more than 1000 times higher. Successful live virus isolation from throat swabs is another striking difference from SARS, for which such isolation was rarely successful^16-18^. Altogether, this suggests active virus replication in upper respiratory tract tissues, where only minimal ACE-2 expression is found and SARS-CoV is therefore not thought to replicate^19^. At the same time, the concurrent use of ACE-2 as a receptor by SARS-CoV and SARS-CoV-2 corresponds to a highly similar excretion kinetic in sputum, with active replication in the lung. SARS-CoV was found in sputum at mean concentrations of 1.2-2.8×10^6^ copies per mL, which corresponds to observations made here^13^.

Whereas proof of replication by histopathology is awaited, extended tissue tropism of SARS-CoV-2 with replication in the throat is strongly supported by our studies of sgRNA-transcribing cells in throat swab samples, particularly during the first 5 days of symptoms. Striking additional evidence for independent replication in the throat is provided by sequence findings in one patient who consistently showed a distinct virus in her throat as opposed to the lung. Critically, the majority of patients in the present study seemed to be already beyond their shedding peak in upper respiratory tract samples when first tested, while shedding of infectious virus in sputum continued through the first week of symptoms. Together, these findings suggest a more efficient transmission of SARS-CoV-2 than SARS-CoV through active pharyngeal viral shedding at a time when symptoms are still mild and typical of upper respiratory tract infection. Later in the disease, COVID-19 then resembles SARS in terms of replication in the lower respiratory tract. Of note, the two patients who showed some symptoms of lung affection showed a prolonged viral load in sputum. Studies should look at the prognostic value of an increase of viral load beyond the end of week 1, potentially indicating aggravation of symptoms.

One of the most interesting hypotheses to explain a potential extension of tropism to the throat is the presence of a polybasic furin-type cleavage site at the S1-S2 junction in the SARS-CoV-2 spike protein that is not present in SARS-CoV^17^. Insertion of a polybasic cleavage site in the S1-S2 region in SARS-CoV was shown to lead to a moderate but discernible gain of fusion activity that might result in increased viral entry in tissues with low density of ACE2 expression^20^.

The combination of very high virus RNA concentrations and occasional detection of sgRNA-containing cells in stool indicate active replication in the gastrointestinal tract. Active replication is also suggested by a much higher detection rate as compared to MERS-coronavirus, for which we found stool-associated RNA in only 14.6% samples in 37 patients hospitalized in Riyadh, Saudi Arabia^21,22^. If virus was only passively present in stool, such as after swallowing respiratory secretions, similar detection rates as for MERS-CoV would be expected. Replication in the gastrointestinal tract is also supported by analogy with SARS-CoV, which was regularly excreted in stool, from which it could be isolated in cell culture^23^. Our failure to isolate live SARS-CoV-2 from stool may be due to the mild courses of cases, with only one case showing intermittent diarrhea. In China, diarrhea has been seen in only 2 of 99 cases^24^. Further studies should therefore address whether SARS-CoV-2 shed in stool is rendered non-infectious though contact with the gut environment. Our initial results suggest that measures to contain viral spread should aim at droplet-, rather than fomite-based transmission.

The prolonged viral shedding in sputum is relevant not only for hospital infection control, but also for discharge management. In a situation characterized by limited capacity of hospital beds in infectious diseases wards, there is pressure for early discharge following treatment. Based on the present findings, early discharge with ensuing home isolation could be chosen for patients who are beyond day 10 of symptoms with less than 100,000 viral RNA copies per ml of sputum. Both criteria predict that there is little residual risk of infectivity, based on cell culture.

The serological courses of all patients suggest a timing of seroconversion similar to or slightly earlier than in SARS-CoV infection^18^. Seroconversion in most cases of SARS occurred during the second week of symptoms. As in SARS and MERS, IgM was not detected significantly earlier than IgG in immunofluorescence, which might in part be due to technical reasons as the higher avidity of IgG antibodies outcompetes IgM for viral epitopes in the assay. IgG depletion can only partially alleviate this effect. Because IFA is a labor-intensive method, ELISA tests should be developed as a screening test. Neutralization testing is necessary to rule out cross-reactive antibodies directed against endemic human coronaviruses. Based on frequently low neutralizing antibody titers observed in coronavirus infection ^12,25^, we have here developed a particularly sensitive plaque reduction neutralization assay. Considering the titers observed, a simpler microneutralization test format is likely to provide sufficient sensitivity in routine application and population studies.

When aligned to viral load courses, it seems there is no abrupt virus elimination at the time of seroconversion. Rather, seroconversion early in week 2 coincides with a slow but steady decline of sputum viral load. Whether certain properties such as glycosylation pattern at critical sites of the glycoprotein play a role in the attenuation of neutralizing antibody response needs further clarification. In any case, vaccine approaches targeting mainly the induction of antibody responses should aim to induce particularly strong antibody responses in order to be effective.

## Data Availability

All data is available in the manuscript and the Annex.

## Materials and Methods

### Clinical samples and viral load conversion

Sputum- and stool samples were taken and shipped in native condition. Oro- and nasopharyngeal throat swabs were preserved in 3 mL of viral transport medium. Viral loads in sputum samples were projected to RNA copies per mL, in stool to copies per g, and in throat swabs to copies per 3 mL, assuming that all sample components were suspended in the 3 mL viral transport medium. For swab samples suspended in less than 3 mL viral transport medium, this conversion was adapted to represent copies per whole swab. An aggregated overview of samples received per day post onset of disease from all patients is shown in **Figure 1A**.

### RT-PCR for SARS-CoV-2 and other respiratory viruses

RT-PCR used targets in the E- and RdRp genes as described^10^. Both laboratories used a pre-formulated oligonucleotide mixture (Tib-Molbiol, Berlin, Germany) to make laboratory procedures more reproducible. All patients were also tested for other respiratory viruses, including human coronaviruses (HCoV) -HKU1, -OC43, -NL63, -229E; Influenza virus A and B, Rhinovirus, Enterovirus, Respiratory syncytial virus, Human Parainfluenza virus 1-4, Human metapneumovirus, Adenovirus, and Human bocavirus using LightMix-Modular Assays (Roche, Penzberg, Germany). Additional technical details are provided in Section 1 in the Supplementary Appendix.

### Virus isolation

Virus isolation was done in two laboratories on Vero E6 cells. 100 µl of suspended, cleared, and filtered clinical sample was mixed with an equal volume of cell culture medium. Supernatant was harvested after 0, 1, 3, and 5 days and used in RT-PCR analysis. Additional technical details are provided in Section 2a in the Supplementary Appendix.

### Serology

We performed recombinant immunofluorescence assays to determine the specific reactivity against recombinant spike proteins in VeroB4 cells, as described^11,12^. This assay used cloned CoV spike protein from HCoV-229E, HCoV-NL63, HCoV-OC43, HCoV-HKU1, and SARS-CoV-2. The screening dilution was 1:10. Plaque reduction neutralization tests were done essentially as previously described for MERS-CoV^12^. Serum dilutions causing plaque reductions of 90% (PRNT90) and 50% (PRNT50) were recorded as titers. Additional technical details are provided in Section 2b and 2c in the Supplementary Appendix.

### Statistical Analyses

Statistical analyses were done using SPSS software (Version 25) or GrapPad Prism (Version 8).

### Ethical approval statement

All patients provided informed consent to the use of their data and clinical samples for the purposes of the present study. Institutional review board clearance for the scientific use of patient data has been granted to the treating institution by the Ethikkommission bei der Medizinischen Fakultät der Ludwig Maximillians Universtität München.

## Acknowledgements

This work was funded by grants from the German Ministry of Research (01KI1723A) and the European Union (602525) to C. D. as well as by the German Bundeswehr Medical Service Biodefense Research Program. The funders had no role in study design, data collection and analysis or decision to publish.

We thank Petra Mackeldanz, Elisabeth Möncke-Buchner, Anja Richter, Marie Schmidt, and Jörn Beheim-Schwarzbach for technical assistance.

## Author contributions

Roman Wölfel: Planned and supervised laboratory testing and evaluated data.

Victor M. Corman: Planned and supervised laboratory testing and evaluated data.

Wolfgang Guggemos: Managed patients and evaluated clinical data.

Michael Seilmaier: Managed patients and evaluated clinical data.

Sabine Zange: Performed laboratory testing.

Marcel A. Müller: Managed serological laboratory testing.

Daniela Niemeyer: Managed and performed virus isolation studies

Patrick Vollmar: Managed laboratory testing

Camilla Rothe: Managed initial patient contacts

Michael Hoelscher: Managed initial patient contacts and evaluated clinical data

Tobias Bleicker: Performed laboratory testing

Sebastian Brünink: Performed laboratory testing

Julia Schneider: Performed laboratory testing

Rosina Ehmann: Performed laboratory testing

Katrin Zwirglmaier: Performed laboratory testing

Christian Drosten: Designed and supervised laboratory studies, wrote the manuscript

Clemens Wendtner: Designed and supervised clinical management and clinical data

## Supplementary Information

The authors declare no competing interests.

